# A Systematic Review on Psychological and Biological Mediators Between Adversity and Psychosis: Potential Targets for Treatment

**DOI:** 10.1101/2019.12.14.19014506

**Authors:** Luis Alameda, Victoria Rodriguez, Ewan Carr, Monica Aas, Giulia Trotta, Paolo Marino, Natasha Vorontsova, Andrés Herane-Vives, Edoardo Spinazzola, Marta Di Forti, Craig Morgan, Robin M Murray

## Abstract

Various psychological and biological pathways have been proposed as mediators between childhood adverse events (CA) and psychosis. A systematic review of the evidence in this domain is needed. The aim of this work is to systematically review the evidence on psychological and biological mediators between CA and psychosis across the psychosis spectrum. This systematic review followed Preferred Reporting Items for Systematic Reviews and Meta-Analyses (PRISMA) guidelines (registration number: CRD42018100846). Articles published between 1979 and July 2019 were identified through a literature search in OVID (PsychINFO; Medline and Embase). The evidence by each analysis and each study results are presented by group of mediator categories found in the review. The percentage of total effect mediated was calculated. 47 studies were included, with a total of 79,668 from general population (GP) and 3,189 from clinical samples. The quality of studies was judged as “fair”. Our results showed (i) solid evidence of mediation between CA and psychosis by negative cognitive schemas about the self, the world, and others (NS); by dissociation and other PTSD symptoms; (ii) evidence of al mediation through an affective pathway (affective dysregulation, anxiety, and depression) in GP; (iii) lack of studies exploring biological mediators. To conclude, we found evidence suggesting that various overlapping and not competing pathways contribute partially to the link between adversity and psychosis. Experiences of adversity, along with relevant mediators such as PTSD and mood related symptoms and NS, should be routinely assessed in patients with psychosis. Targeting such mediators through cognitive behavioural aproaches using trauma-focused therapy and/or pharmacological means could be a useful addition to the traditional treatment of positive symptoms.

## Introduction

Evidence has accumulated over the past 15 years showing that exposure to childhood adversity (CA) – in the form of abuse, neglect and bullying – is associated with increased risk of psychosis across the spectrum, from low-level experiences to disorder^1^. Moreover, many studies have demonstrated that patients already suffering from psychosis who were exposed to CA experience higher severity of more severe positive symptoms, particularly hallucinations and delusions, compared with their non-traumatised counterparts^2, 3^. However, research to date is less clear on the mechanism involved. Therefore, a systematic review of the evidence about possible mediating mechanisms linking CA and psychosis is needed.

In terms of biology, substantial research on the underlying biological mechanisms of the CA-psychosis association has been carried out in the last 15 years. This covers dysfunction in pathways such as the stress response system^4^, dopaminergic neurotransmission^5^, inflammation and redox dysregulation^6^, and changes in stress related brain structures such as the amygdala or the hippocampus^7^. For example, it has been suggested that excessive exposure to stress might lead to an overactivation of the Hypothalamic-Pituitary-Adrenal (HPA) Axis. This could be toxic for hippocampal functioning ^8 9^, which might in turn contribute to the emergence of psychosis. Furthermore, acute social stress has also been found to increase striatal dopamine release in individuals at risk for psychosis, in patients with psychotic disorder, and in individuals with childhood trauma ^5^. In addition, inflammatory dysfunctions as well as redox dysregulation conditions^6, 10^ have been found among individuals exposed to CA and among patients with psychosis.

Several psychological models have also been proposed to explain the relationship between CA and psychosis. These models look at the same epiphenomena from different angles and can be complementary, thus they should not be considered necessarily as competing explanations. One theory postulates that CA may lead to psychosis through a pathway of heightened emotional distress, characterized by hypersensitivity to daily-life stressors, leading to anxiety and depression^11, 12^ and it is often called “Affective Pathway to Psychosis”. These symptoms, sometimes called “ancillary symptoms of psychosis” ^12^ might operate as mediators between adversity and positive symptoms of psychosis. Another model proposes that severe forms of adversity might lead to cognitive biases, such as negative schema about the self and the world, which, in addition to a heightened tendency to attribute experiences to external causes, might give rise to paranoia, ideas of reference and misattributions of perceptual abnormalities^13, 14^. A third explanatory model relies on Attachment Theory^15^, which hypothesizes that adversity occurring within the context of caregiving relationships might disrupt the quality of attachment styles leading to insecure, fearful or anxious attachment. This might, in turn, play a role in psychosis itself, but also in processes contributing to treatment effectiveness, such as therapeutic alliance or engagement with services^16^. Finally, another putative pathway linking adversity with psychosis emphasises the mediating role of Post Traumatic Stress Disorder (PTSD) and related symptoms, such as dissociation, depersonalisation and intrusive memories, very frequently comorbid with psychotic phenomena^17^. For example, It is suggested that these experiences could be interpreted as being externally generated, leading to hallucinatory experiences and hampering reality testing^13, 18^.

Despite several narrative reviews covering potential biological pathways and psychological mediating pathways^7, 12, 19-23^, to date only a single systematic review has been conducted^24^. Williams et al.,^24^ examined psychological mediators between CA and psychosis including papers up to September 2017, which consisted of a total of 37 studies. They found robust evidence for a mediation by cognitive bias, such as negative schemas about the self and the world, by symptoms of PTSD, and by affective processes. However, this review is limited by the fact that (I) they did not analyze and discuss the evidence of all the mediation pathways tested within each study and limited to the summary of the main conclusions provided by each author; (ii) no summary was given regarding the amount of mediation between adversity and psychosis (e.g. no information on the percentage of total effect mediated was systematically extracted and summarized) (iii) their review did not include biological studies.

The present study overcomes these limitations and includes additional papers published up to July 2019, providing an additional period of 22 months of research in the field. Our aim is to systematically review the evidence on potential psychological and biological mediators between CA and psychosis (considered as presenting hallucinations, delusions or disorganisation) across the spectrum from low level experiences in general populations to disorder. Results will be grouped based on existing proposed theoretical mechanisms and discussed in terms of potential treatment interventions.

## Methods

### Search strategy

A systematic review of the literature was conducted following Preferred Reporting Items for Systematic Reviews and Meta-Analyses PRISMA^25^ guidelines. The systematic review was registered in PROSPERO^26^ in July 2018 (registration number: CRD42018100846). The main search was conducted on MEDLINE, EMBASE and PsycINFO, through Ovid provider in June 2018 and it was updated in July 2019. We searched Medical Subjects Headings (MeSH) and keywords related to: (1) childhood adversity, including exposure to sexual, physical, emotional abuse, physical and emotional neglect; bullying and early separation from parents; (2) mediation and (3) psychosis, using the Boolean operator ‘AND’ (full list of search terms provided in **Online Supplementary Material –** *Search Strategy*). The present analyses did not consider separation from parents, abandonment and parental loss given the heterogenous definitions used to describe such type of adversities across studies, in keeping with others^1^.

Titles and abstracts of articles were screened independently by two reviewers (LA and VR) with 95% and 85% agreement (agreement was considered when equal quality assessment scores were) for articles on psychological and biological mediators, respectively. Discrepancies were resolved through discussion at a project group meeting.

### Inclusion and exclusion criteria

Included studies were those that (1) examined psychological or biological mediators of the relationship between CA and (i) psychosis onset, (ii) severity of positive symptoms in patients with a psychotic disorder, or (iii) severity of attenuated psychotic symptoms in the general population; (2) employed a robust method for testing mediation in the analyses and fulfilling the Baron and Kenny criteria^27^; (3) for clinical samples studies only (not for general population), psychotic disorder was defined according to Diagnostic and Statistical Manual of Mental Disorders (DSM-III, DSM-III-R, DSM-IV, DSM IV-TR^28^), Research Diagnostic Criteria or International Classification of Diseases, Ninth or Ten Revision (ICD-9), (ICD-10)^29^; (4) assessed both mediators and outcomes using validated methods and scales; (5) included CA occurring before age 18 and involving exposure to sexual, physical and emotional abuse, physical and emotional neglect, bullying or equivalent experiences; (6) had been published as original research; and (7) were performed in humans.

Exclusion criteria were: (1) not being published in English language; (2) including more than 20% of participants aged 65 years or over, in accord with others^30^; (3) being performed in homogeneous samples of specific populations such as pregnant women or samples from forensic settings; (4) including non-psychological mediators (e.g. being exposed to later life hassles, level of education, different aspects of social disadvantage or drug use were not included in the current work despite of the potential of being targeted by social or motivational interventions).

### Quality Assessment and data extraction methodology

Quality assessment was carried out using the Newcastle–Ottawa Scale^31^ for cohort studies by two independent reviewers (LA and PM). Details on the instrument items and quality assessment procedures can be found in **Supplementary material section**. The agreed quality grades of each study are presented in **Supplementary Material STable 1a**, 1**b** and 1**c**.

**Table 1.** Summary of evidence for mediators between adversity and psychosis in clinical samples (subjects at risk for psychosis and with psychotic disorder) and in general population

| **Category** | **Clinical samples** |  |  | **General population** |  |  | **Total** |
| --- | --- | --- | --- | --- | --- | --- | --- |
|  | *Number of analyses.*<br>*(number of studies)* |  |  | *Number of analyses.*<br>*(number of studies)* |  |  |  |
| **(N total analyses/studies\*)** | Evidence of mediation | Null mediation | % analyses supporting mediation | Evidence of mediation | Null mediation | % analyses supporting mediation | % analyses supporting mediation |
| **Dissociation (43/12)** | 6(4) | 20(6) | **23%** | 12(6) | 5(1) | **70%** | **42%** |
| **Dysfunctional attachment (18/5)** | 1(1) | 3(2) | **25%** | 7(3) | 7(2) | **50%** | **44%** |
| **Affective pathway (32/14<sup>ab</sup>)** | - | 3(1) | **0%** | 21(9) | 8(5) | **72%** | **66%** |
| **Loneliness (7/4)** | 1(1) | - | **100%** | 4(3) | 2(1) | **66%** | **83%** |
| **Cognitive schemas (53/20<sup>b</sup>)** | 8(6) | 13(5) | **38%** | 20(8) | 12(6) | **62%** | **53%** |
| **Others PTSD symptoms (7/4)** | 5(3) | 1(1) | **83%** | 1(1) | - | **100%** | **86%** |
| **Others<sup>c</sup> (10/6)** | 1(1) | 1(1) | **50%** | 7(3) | 1(1) | **87%** | **80%** |

<sup>a</sup>Isvoranu et al., was only included in the articles count within but not in the analyses count. <sup>b</sup>Ashford et al., was only included in the articles count but not in the analyses count. <sup>c</sup>Others included anomalous self-experiences; mania; time-perspective capabilities and compulsions as mediators. \* We considered that one analysis showed evidence of mediation when authors reported significant p-values (<0.05) in the indirect (or mediating) effects, or when using a regression-based approach, an important reduction of the total effect occurred once the mediator was included in the model. Null mediation was defined as the non significant (p>0.05) indirect or mediating effect or as the lack of reduction of the total effect once the mediator was included in the model when using a regression-based approach.

To summarise the evidence of mediation between adversity and psychosis across the spectrum (from low level psychotic symptoms in the general population, to attenuated psychotic symptom in subjects at risk for psychosis and in patients suffering from the disorder) we constructed Tables **1, 2, 3** and **STable 2**. We enumerated the number of studies and analyses indicating whether there was evidence for mediation or lack of mediation in clinical samples (including subjects at risk and with psychotic disorder) and in general population studies. These results were extracted from the text or tables of each paper. We included all mediation analyses (i.e. distinct pathways tested) from each paper, meaning the total number of analyses was greater than the number of papers. In **Tabes 1, 2, 3** and **STable 2**, as well as in the text we provided the “percentage of analyses showing evidence of mediation”, across all categories of mediators. We considered that one analysis showed evidence of mediation when authors reported significant p-values (<0.05) in the indirect (or mediating) effects, or when using a regression-based approach, an important reduction of the total effect occurred once the mediator was included in the model. This allowed us to provide the percentage of analyses supporting mediation per category (as an example: for dissociation in general population there were 17 pathways tested across 6 different studies, 12 of them were supportive of mediation and 5 were not, thus 70% (12/17) of analyses were supportive of mediation through dissociation in the general population).

**Table 2.** Summary of evidence for mediators within the Affective Pathway category between adversity and psychosis in clinical samples (subjects at risk for psychosis and with psychotic disorder) and in general population

|  | Clinical samples |  |  | General population |  |  | Total |
| --- | --- | --- | --- | --- | --- | --- | --- |
| Category | Number of analyses.<br>(number of studies) |  |  | Number of analyses.<br>(number of studies) |  |  |  |
| (N total analyses/studies) | Evidence of mediation | Null mediation | % analyses supporting mediation | Evidence of mediation | Null mediation | % analyses supporting mediation | % analyses supporting mediation |
| **Affective pathway (32/14)<sup>ab</sup>** | - | 3(1) | **0%** | 21(9) | 8(5) | **72%** | **66%** |
| **Anxiety (7/4)** | - | 1(1) | **0%** | 4() | 2(2) | **66%** | **57%** |
| **Depression (10/6)** | - | 1(1) | **0%** | 4(2) | 4(3) | **55%** | **55.5%** |
| **Emotional dysregulation (8/4)** | - | 1(1) | **0%** | 5(2) | 2(2) | **71%** | **62.5%** |
| **Stress sensitivity/perception (5/2)** | - | - | - | 5(3) | - | **100%** | **100%** |
| **Composite (2/1)** | - | - | - | 5(2) | - | **100%** | **100%** |

<sup>a</sup>Isvoranu et al., was only included in the articles count within but not in the analyses count. <sup>b</sup>Ashford et al., was only included in the articles count but not in the analyses count.

Given that considering mediation based on the significance of the p-value is highly dependent on the sample size and thus very limited, we also collected information on the percentage of total effect that was mediated, which is equivalent of the amount of mediation in each pathway tested. **Figure 2** represents the percentage of the total effect mediated by each analysis for which information was available across the most meaningful categories described in the results section. The type of adversity and outcome are also shown to have a visual representation of the pathway. As an indicative measure, the median value of all the analyses per category is also highlighted in the figure and presented in the text. Where the “proportion of total effect mediated” was not given by authors, we derived an estimate by dividing the indirect effect by the total effect and multiplying it by 100 in keeping by others^32^. Further details on data extraction procedures can be found in **Supplementary materials** and in **Figure 2** footnote. In **STables 1a, 1b** and **1c** we also provide details on the significance of the indirect and direct effects as well as the proportion of total effect mediated in each study.

**Figure 1.**
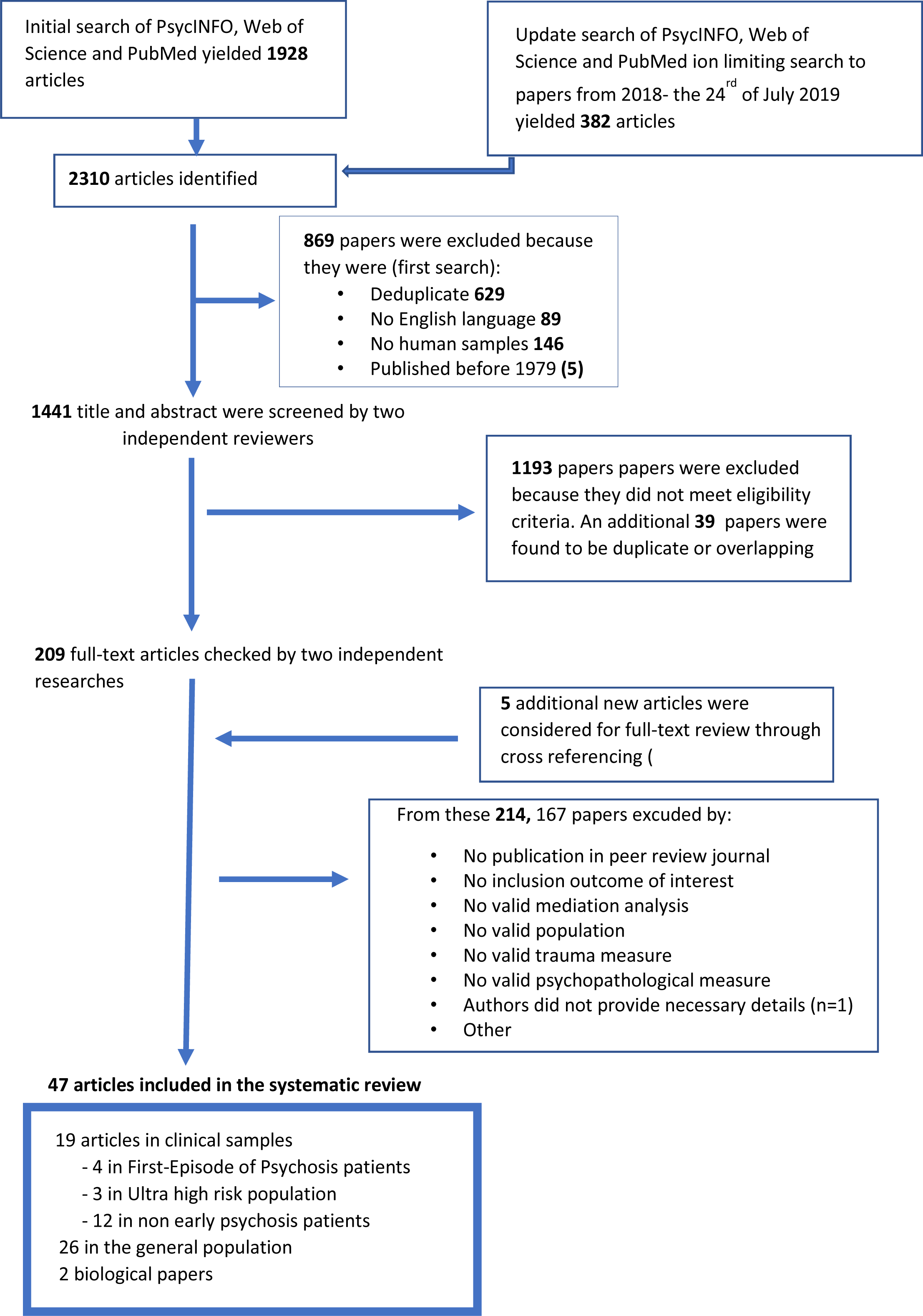
Flow chart.

**Figure 2.**
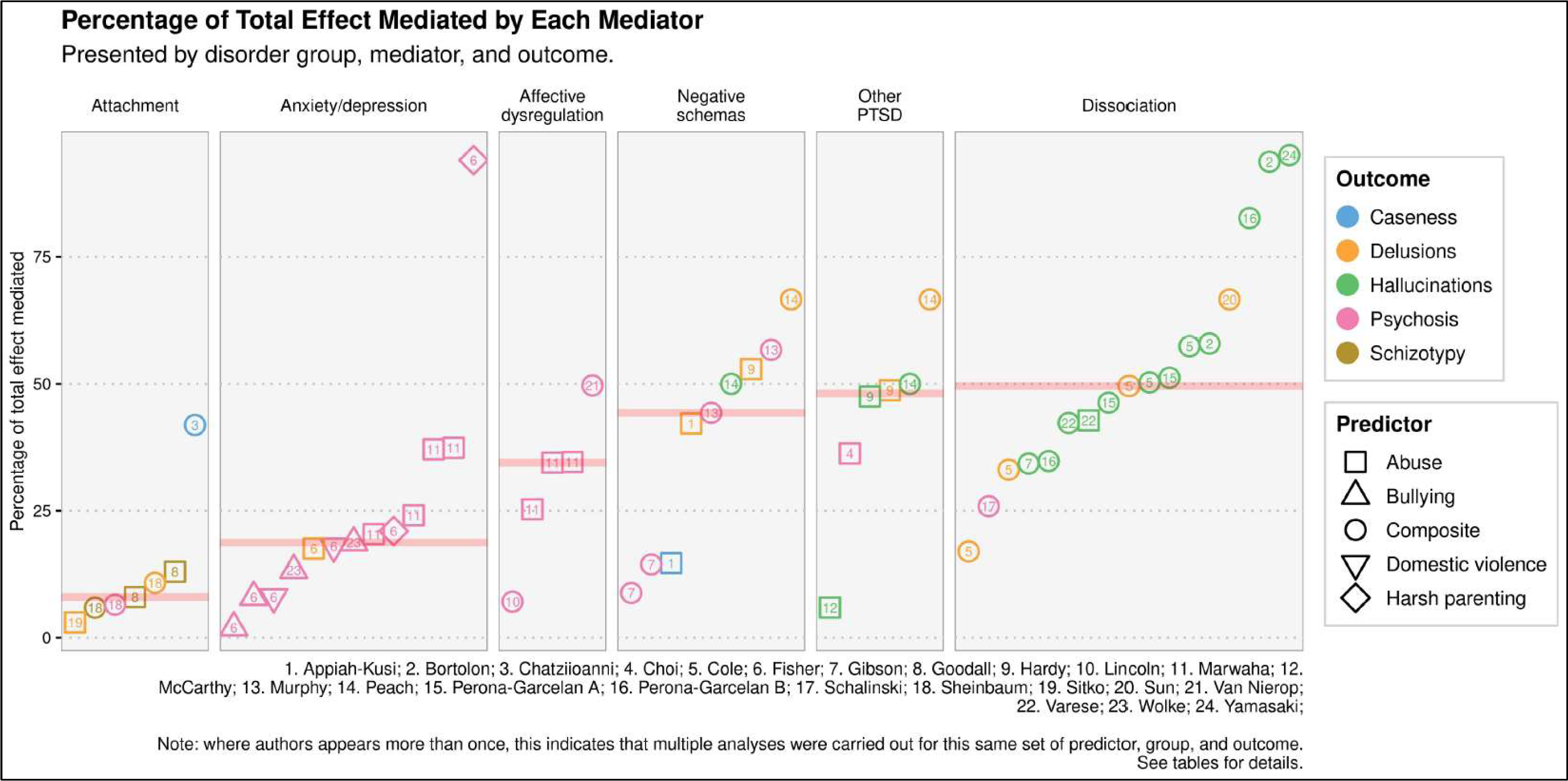
We managed to obtain the percentage of total effect mediated for for 83.1% of the analyses that found a significant indirect effect (103 out of the 124 positive analyses, 44 of which were already calculated and 59 which were calculated as described in **Supplementary Material** - Data extraction procedures section). In this table are presented the percentage of the most meaningful categories of mediators found in our results. As presented in the right-hand side of the figure, the type of predictor is detailed by different symbols and the type of outcome is detailed by different colours.

## Results

We identified 47 studies that met our inclusion criteria from the 2310 studies found in the initial searches (2018 and 2019 combined) (Figure 1).

Twenty-one studies were conducted in clinical samples (four in First Episode of Psychosis (FEP) or early psychosis patients ^33-36^, three in Ultra High Risk (UHR) patients^37-39^, 14 in non-FEP patients^40-53^ 26^54-79^ in the general population. Our review included 79,668 subjects from the general population and 3,189 subjects from clinical studies. Participant ages ranged from 18.5 to 44.6 years old in clinical samples (30.4 in average) and between 9.8 to 51.7 in the general population (34.3 in average). In clinical samples, 35% of the participants were women, compared with 37% of the volunteers.

This total number of analyses excludes two studies with an extremely high number of analyses ((one used a network based approach exploring multiple connections between adversity and symptoms^80^, the other included up to 28^54^). These studies would have distorted the numerical summaries and therefore are described in the text only. Overall, 170 analyses were included in this review (ranging between 1 and 12 analysis per paper).

The quality check agreement between the two raters was 81.8%. Overall, quality was graded as ‘fair’ (between 4 and 7) for all studies except one, where quality was judged as ‘good^60^. Studies of biological mediators tended to be graded with lower scores failing to provide estimates of the indirect effects, relying on small samples, and/or cross-sectional data^40, 41^. Overall, only 4/47 studies^39, 60, 65, 77^ were used a prospective design. Eight^41, 54^-^57, 64, 67, 80^ of the 47 studies reported a mediating effect but failed to provide estimates of the indirect and direct effects and thus were classified as ‘suggested mediation’, as described above.

**Table 1** summarises the evidence for each analysis by mediator category. A comprehensive table containing all extracted data including quality scores is available as online **Supplementary material (STable 1a, 1b, 1c)**. Given the high number of analyses and the heterogeneity found for the affective pathway, cognitive schema pathway, and dissociation, three detailed tables were constructed for these groups (see **Table 2, Table 3** and S**Table 2** in **Supplementary material** respectively) presenting the evidence for each subcategory.

**Table 3.** Summary of evidence for mediators within the Cognitive Schemas category between adversity and psychosis in clinical samples (subjects at risk for psychosis and with psychotic disorder) and in general population

|  | Clinical samples |  |  | General population |  |  | Total |
| --- | --- | --- | --- | --- | --- | --- | --- |
| Category | Number of analyses.<br>(number of studies) |  |  | Number of analyses.<br>(number of studies) |  |  |  |
| (N total analyses/studies) | Evidence of mediation | Null mediation | % analyses supporting mediation | Evidence of mediation | Null mediation | % analyses supporting mediation | % analyses supporting mediation |
| **Cognitive schemas (53/21)<sup>b</sup>** | 8(6) | 13(5) | **38%** | 20(8) | 12(7) | **62%** | **53%** |
| **Negative schemas (16/7)** | 5(3) | 2(1) | **71%** | 5(4) | 4(1) | **55%** | **62.5%** |
| **ELC (5/3)** | - | - | - | 4(2) | 1(1) | **80%** | **80%** |

<sup>b</sup>Ashford et al., was only included in the articles count but not in the analyses count.

The proportion of total effect mediated could be obtained for 103 out the 124 analyses showing evidence of mediation. **Figure 2** summarises the percentage of total effect mediated for 57 analyses included in 24 papers across the most relevant category of mediators found in our review (only categories being explored in at least three papers are shown in **Figure 2**, the percentage of total effect mediated of the remaining analyses are shown in **STables 1a, 1b** and **1c**).

The most relevant categories of mediators found in our review were: Dissociation (42 analyses, 12 papers), dysfunctional attachment (18 analyses, 5 papers), affective pathway to psychosis (32 analyses, 14 papers), loneliness (6 analyses, 5 papers), cognitive schemas (53 analyses, 20 papers), other PTSD symptoms (7 analyses, 4 papers), other psychological mediators (10 analyses, 6 papers), biological mediators (2 analyses, 2 papers).

As a whole, in terms of amount of mediation between CA and psychosis and the percentage of analyses showing evidence of mediation across all categories, we found evidence suggesting that various overlapping pathways contribute partially to the link between adversity and psychosis. Specifically we found (i) solid evidence of mediation between CA and psychosis by negative cognitive schemas about the self, the world and others as well as solid evidence of mediation by PTSD symptoms, particularly dissociation. We found evidence of mediation through an affective pathway (particularly for affective dysregulation, anxiety and depression) in the general population with no evidence of mediation in clinical samples. Although only explored in a limited number of studies, the feeling of loneliness appeared to partially mediate the link between CA and psychosis. The was no evidence supporting mediating role of the remaining categories.

### Dissociation

Despite only 43% of analyses showing evidence of mediation, 10/12 studies^36, 55, 58, 62, 70, 72, 78, 79, 81-83^ were supportive of mediation, against 2/12 that were not^33, 84^. The percentage of total effect mediated (which is the amount of the adversity-psychosis association that is explained by the mediator – see methods and **Figure 2** footnote for details) was consistently high across analyses within this category, with a median of around 50% of total effect explained. Moreover, as illustrated in **Figure 2**, most of the studies showing evidence of mediating effects used hallucinations as outcome and explored trauma as a composite score^55, 58, 62, 70, 78, 82, 83^, while only 2^36, 58^ studies found positive effects with delusions as the outcome (see also **STables 1a** and **1b** for detail**s**). As shown in **STable 2**, due to high levels of heterogeneity for specific items within the dissociative category (depersonalisation, absorption, dissociative amnesia and defensive dissociation), it was not possible to draw conclusions about which dissociative symptoms were more likely to mediate the adversity-psychosis association.

### Dysfunctional attachment

Despite overall 43% of analyses showing evidence of mediation, 3/5^42, 73, 75^ papers showed mixed findings, 1/5^85^ showed no evidence of mediation and just 1/5^63^ showed consistent evidence of mediation across all its analyses. As seen in **Figure 2**, the percentages of total effect mediated were the lowest among all categories, with a median value of around 12% of effect mediated. Studies examining different outcomes and types of traumas were not enough to allow conclusions about specific mediational pathways.

### Affective Pathway

Overall, only one paper explored this pathway in clinical samples^39^ showing no evidence of mediation. The remaining 13^54, 59, 60, 62, 65-68, 72, 76^-^78, 80^ papers (including 29 analyses) were conducted in general population samples, with 66% of analyses supporting mediation (Ashford et al.,^54^ and Isvoranu et al.,^80^ not included in the analyses count as mentioned above). 11/13^54, 59, 60, 62, 65^-^67, 72, 76, 77, 80^ of these general population studies found evidence of mediation and 3 of them were prospective graded high in quality assessment^60, 65, 77^ **STable 1b**).

As mentioned before, we divided the Affective Pathway into the following discrete subcategories: Anxiety, Depression, Affective dysregulation, Stress sensitivity and a residual composite measure (details on the direction of evidence can be found in **Table 2)**. Briefly, in terms of proportion of analyses supporting mediation, results per subcategories showed that 57% of analyses for anxiety, 55% for depression, 62% for affective dysregulation and 100% for stress sensitivity showed evidence of mediation. As seen in **Figure 2**, the median percentages of total effect mediated (the amount of the adversity-psychosis association that is explained by the mediator) for anxiety and depression were at around 20%; while for affective dysregulation’s they rose up to approximatively 35%. As illustrated in **Figure 2**, the majority of studies examined a composite category of attenuated psychotic symptoms as outcome and tended to examine specific types of adversities such as abuse and bullying.

### Feeling of Loneliness

83% of analyses showed evidence of mediation. Only one study^48^ was conducted in clinical samples and showed evidence for mediation. In general population, two studies showed evidence of mediation^56, 64^ and another showed mixed findings^74^. The low number of analyses did not allow us to draw consistent conclusions in terms of specific pathways between adversity-psychosis (see **STable 1a, 1b** for details).

#### Cognitive schemas

53% of analyses overall showed evidence of mediation in this category. In terms of papers, 15/20 were supportive of mediation ^33, 35, 37, 38, 54, 55, 57, 60, 62, 64, 69, 71, 76, 86, 87^, 5/20 were not^34, 59, 70, 78, 88^. Results were more likely to show evidence of mediation when conducted in the general population, compared with clinical samples (65% versus 38% of analyses were supportive of mediation respectively, see **Table 1**).

As displayed in **Table 3**, we divided this category into: Negative schemas about self, others and the world and External Locus of control (ELC). Briefly, the evidence for the former showed that 62.5% of analyses were supportive of mediation); with a proportion of total effect mediated (the amount of the adversity-psychosis association that is explained by the mediator) of around 47% (median value across analyses in this category – see **Figure 2**). No studies in clinical samples explored the ELC and 2.3 studies in general population showed evidence of mediation^60, 62^. Other biases such as maladaptive schemas^55, 57^ or low self-steem^60^ showed interesting findings but were not explored specifically due to insufficient number of studies to draw conclusions. In terms of specific adversities and outcomes, studies were very heterogeneous thus not allowing us to draw conclusions about specific pathways.

### Other PTSD symptoms

These included posttraumatic intrusions, avoidance, and numbing, or a general measure of posttraumatic symptoms. Overall, results suggested evidence of mediation^35, 68, 87, 89^. The percentage of total effect mediated (the amount of the adversity-psychosis association that is explained by the mediator) was also high, with a median value just below 50% (see **STables 1a, 1b)**

### Other psychological mediators

Other mediators including 4 studies containing 10 analyses that did not fit into the aforementioned categories were grouped in another group. Mediators such as Anomalous self-experiences^61^ and Time perspective capabilities^49^ found promising results, while mania showed no mediation effects^39^. MacCarthy-Jones et al.,^68^ showed that while there was a positive mediating effect for compulsions, this was not the case for obsessions (see **STable 1a, 1b)**.

### Biological mediators

Surprisingly, only 2 studies examining biological mechanism as potential mediators between CA and psychosis fulfilled the inclusion criteria for this systematic review (**STable 1c)**. One study found no evidence for a mediation of the inferior frontal gyrus activation between CA and the Positive and Negative Syndrome Scale (PANSS) positive scores^40^; another showed that the grey matter density in Dorsolateral prefrontal Cortex (DLPFC) was mediating the link between emotional neglect and disorganized symptoms in patients with psychosis^41^.

## Discussion

From this systematic review of 47 papers, we found evidence of partial mediation between adversity and psychosis through various overlapping, and not competing, psychological mechanisms. Adversity and psychosis was particularly driven by negative cognitive schemas about the self, the world and others, and by dissociation and other PTSD symptoms. For general population samples there was good evidence for mediation through an affective pathway (affective dysregulation, anxiety and depression), but there were insufficient studies looking at this in individual at risk for psychosis and in patients suffering from the disorder to allow conclusions to be drawn about its mediating role in clinical settings.

Our systematic review found no evidence supporting the mediating role of dysfunctional attachment styles in the general population; there were too few studies addressing this mediator in individual at risk and in subjects with the disorder to permit conclusions. Other mediators showing interesting findings included loneliness, stress sensitivity, and external locus of control, but generalisation of findings is limited due to the low number of studies. However, they should be taken into account in future research. Contrary to our expectations, only two papers^40, 41^ fulfilled our inclusion criteria examining biological mediators. There is evidence that potential biological mediators are associated with adversity and with psychosis, but very few considering mediation effects directly. So, the speculation on biological mediators is currently mostly conjecture showing an urgent need for more research in this field.

## Strengths and limitations

The findings of this review should be interpreted in the context of various strengths and limitations. A major strength is the large scale of this review including 47 studies, 79,668 subjects from the general population and 3,189 from clinal studies. This has allowed us to cover multiple mediator groups with a sufficient number of participants and show new pathways that did not appear in William et al., systematic review^24^, such as the role of loneliness. Second, we have not limited our analysis to the description of the main finding of each study and to report whether the mediation was present or absent, but we have also examined the amount of mediation of each pathway by providing the percentage of total effect mediated (when this was possible). We believe this is an important point given that just limiting to the significance of the p-value of the indirect effects is highly dependent on the sample size and thus totally limited. Calculating the proportion of the effect that is mediated provides a more accurate understanding of the mediational processes. Moreover, we believe that our concrete clinical implications may be useful and could contribute to a better knowledge on trauma-informed care in services treating individuals with psychosis.

Despite these positive aspects, some limitations must be mentioned. First, only three studies used a prospective design to estimate the indirect effects, while in the remaining studies it cannot be excluded that the mediator resulted as a consequence of psychosis. Second, CA was measured retrospectively in all studies except two^60, 77^ which were conducted in the general population. In patients this can constitute a risk of bias given the difficulties of patients to recall their experiences and to disclose these openly in assessment by research assistants, which is especially true in people with psychosis. Third, unfortunately the percentage of total effect could not be obtained nor calculated in 17% of the analyses that were supportive of mediation (give that authors did not provide details on the indirect, direct and total effects which are necessary to the calculation), thus our **Figure 2** is not totally representative of the total number of analyses included in this review. In addition, some mediators (i.e. attachment styles, anxiety and depression) were found to be highly explored in the general population but very little in clinical samples, which is an important limitation of current literature in the field and which limits the extent to which we can extrapolate our conclusions to clinical intervention.

Furthermore, papers in this review considered that null mediation occurred when the indirect or mediating effects did not reach a significant p-value of <0.05. As aforementioned, considering the p-value to test the null hypothesis is limited by the fact that is highly dependent on the sample size. Thus, it is likely that in our review studies conducted in small samples are underestimating potential mediating effects, and this should be taken into consideration when interpreting our conclusions.

### Evidence for mediational pathways between childhood adversity and psychosis

#### Pathway 1: Dissociation and PTSD symptoms

Mediation through this pathway was more common when hallucinations were used as an outcome. These two categories, intimately related, showed the highest percentages of total effect mediated (**Figure 2**).

The role of post-traumatic dissociation in psychosis was already the focus of research at the end of the eighteenth century, when Janet, among others, defined Hysterical Psychosis (HP) as characterised by its dissociative and stress related nature^90^. Despite these longstanding connections between dissociation and psychosis, current international classifications do not include dissociation among the diagnostic criteria for any form of psychosis. However, contemporary authors such as Ross^91^ and Moskowitz^90^ suggest the existence of a dissociative type of psychosis that could potentially be responsive to psychotherapy focused on trauma.

Accordingly, our findings showing mediation by other PTSD symptoms in clinical samples^35, 87, 89^, are consistent with reports suggesting that similar mechanisms could be involved in psychotic experiences and symptoms of PTSD. For instance, it has been suggested that some hallucinations represent a dissociated type of posttraumatic intrusion, which may not be recognized as such by people with psychosis^18, 90^. A clinical implication is that if a person experiencing verbal auditory hallucinations recognized the identity or content of a heard voice as corresponding to the content of a previous traumatic event, the intrusion could be reappraised as a type of intrusive memory. It could then be understood in the same way as a re-experiencing symptom of PTSD (instead of an externally attributed voice reflective of psychosis^13^), and could be addressed therapeutically using trauma-focused therapies, such as cognitive behavioural therapy (CBT). Our results therefore support the possibility of applying specific psychological interventions for patients with psychosis, dissociation and PTSD symptoms and with a history of clear-cut traumatic episode. In this regard, two recent systematic reviews exploring the safety and efficacy of trauma focused therapies in individuals with comorbid PTSD symptoms and psychosis have shown that trauma focused CBT is safe^92^ and can reduce PTSD symptoms^93^. Moreover, a meta-analysis of 12 studies also explored its impact on positive symptoms and found small improvements after treatment^94^.

#### Pathway 2: Cognitive schemas

Cognitive schemas about the self, the world and others were the most consistent mediator between CA and psychosis in both clinical samples and general population. Moreover, this category contributed highly to the total effect between CA-psychosis, with median percentage mediated of around 46% across all analyses (**Figure 2**).

This evidence supports previous cognitive models of the development of positive symptoms^13, 95^: these models suggest that exposure to severe trauma might contribute to the development of cognitive bias such as negative schemas about the world, others and the self. Further exposure to stressors or to subtle perceptual abnormalities that are common in the general population^30^, and even more common in genetically predisposed individuals, will lead to anomalous conscious self-experiences which will trigger the search for an explanation. The biased schemas, in combination with anomalous self-experience disrupt the appraisal process leading to a misinterpretation of reality, and subsequently delusional ideas^13, 95, 96^. Other factors are important in this process, such as the tendency to search for an explanation in an external source (otherwise called External locus of control^84^) which although not explored intensively in our review, has shown evidence for mediation, especially in the general population as detailed above^60, 62^. It would be interesting for future research to explore if other cognitive biases such as jumping to conclusions and meta cognitive deficits can moderate or mediate the connexions between CA and psychosis in combination with biases such as negative schemas and External Locus of control.

#### Pathway 3: Affective pathway

We found consistent evidence suggesting that anxiety, depression and affective dysregulation might partially mediate the link between CA and low-level psychotic experiences in the general population. The affective component could be a mediational partner along with other mechanisms such as cognitive bias or PTSD related symptoms. For example, Fisher et al.,^60^ report on a large prospective study, where 100% of the total effect was mediated only when anxiety and depression were added to the model in combination with cognitive bias and low self-esteem.

Unfortunately, our interpretation is restricted to the general population (and thus to attenuated positive symptoms) due to the lack of studies performed in clinical settings, which limits the applicability to possible clinical interventions. Nevertheless, these results support previous claims that the association between social adversity and psychosis might be mediated by non-psychotic symptoms (otherwise the so called “ancillary symptoms of psychosis” ^12^ such as anxiety and depression and affective dysregulation). Stress sensitivity related to daily life activities can lead to negative affect in vulnerable individuals previously exposed to CA, which in turn can affect mood and anxiety. These symptoms might be determinants of paranoid thinking; for example, anxiety might lead to anticipation of threat, and low mood might drive negatively biased interpretations of ongoing experience and impact self-esteem and negative schemas about the self, which in turn are precursors of psychotic symptoms, as illustrated in **Figure 3**.

**Figure 3.**
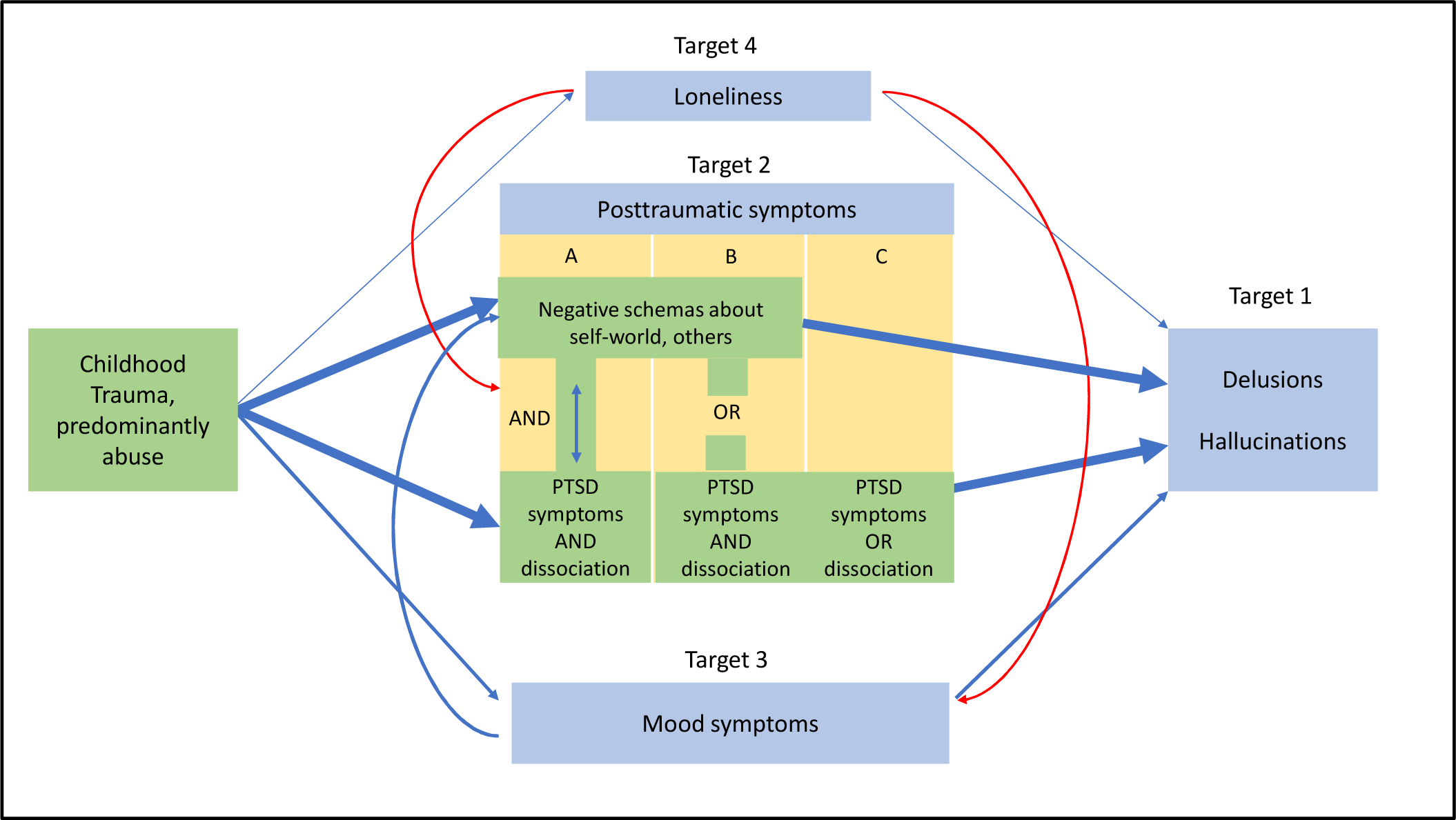
This figure displays the potential targets for treatment that could be addressed in individuals with psychosis who have been exposure to adversity. Beyond the traditional treatment of psychotic symptoms (**Target 1**) different clinical presentations with various targets for treatment can occur: **Target 2** represent targets from the posttraumatic spectrum such as Negative Schemas about the Self, Others and the World (NS) and PTSD related symptoms, including Dissociation. These can be combined in different forms as displayed in **Target 2A, 2B** and **2C. Target 3** represents mood-related symptoms and **Target 4** represent the presence of feeling of loneliness. The treatment options could be adapted depending on which targets are present or predominant in the clinical picture. For **Target 2 A**: CBT-trauma focused therapy (for PTSD symptoms and NS) + sensory grounding techniques (for dissociation) + SSRI/ antagonists α1 if required (for PTSD symptoms). The implementation of these interventions would be adapted according to the presence/absence of the aforementioned targets (**Target 2 B and C**). **Target 3:** CBT techniques such as behavioural activation and graded exposure, relaxation techniques and mindfulness + SSRI if needed for anxiety and depressive symptoms. **Target 4:** Promoting social inclusion and community membership through therapy/activity groups and vocational support. Improving existing interpersonal relationships using family therapy approaches. The thickness of the arrows shows the robustness of the evidence according to our results. The red arrows represent a hypothetical connexion not addressed in our review.

#### Pathway 4: Feeling of Loneliness

Although only explored in 4 studies^48, 56, 64, 74^, we found good evidence suggesting that a feeling of loneliness might mediate the CA-psychosis relationship. We could hypothesize that social withdrawal and loneliness may increase an individual’s sensitivity to potential stressors in daily life restrict access to balanced information from the environment, maintaining biased cognitive schemas. This could, in turn, predispose an individual to lower mood and anxiety, which would constitute a favourable ground for the emergence of psychotic symptoms. Thus, the feeling of loneliness and isolation must be taken very seriously as it might operate as a potential condition allowing other mediators to operate.

#### Pathway 5: Biological measures

Despite the high emphasis put on the search for biomarkers during the last 20 years of psychiatric research, we could identify just two eligible papers exploring biological mechanisms using psychosis as an outcome^40, 41^, which prevents us from drawing conclusions about specific pathways. This highlights the limited evidence in this field and need for future studies addressing different biomarkers comparing between groups of individuals with or without trauma, and to conduct longitudinal designs allowing us to test the influence of biological processes on the onset and evolution of symptoms of psychosis.

### Clinical implications

Our systematic review provides evidence to support some potential treatments for traumatised individuals with psychosis, in addition to the treatment of positive symptoms, which often remain high in this vulnerable group^2, 3^. **Figure 3** displays a model with the different potential treatment targets derived from the pathways mentioned above. Beyond the traditional treatment of psychotic symptoms (Target 1 in **Figure 3**) to which some traumatised patients do not respond, we propose targeting the relevant mediators found in our review, hypothesizing that an improvement in such targets would then have an indirectly beneficial effect on positive symptoms.

A common clinical picture corresponds to a situation where a traumatised patient with psychosis suffers from negative schemas about the self, others and the world and PTSD symptoms, including dissociation. If these are present (Target 2 A in **Figure 3**), a trauma focused therapy, such as trauma-focused cognitive behavioural therapy (CBT), could be appropriate, integrating treatment of negative schemas and traumatic intrusions, in addition to dissociative symptoms. Briefly, this could include grounding techniques, imaginal exposure, memory updating and cognitive restructuring, alongside more general CBT elements such as the use of behavioural experiments to gather new information about current safety^17^. In some cases, dissociative symptoms may be very disruptive and prevent access to the other targets; in this case they can be prioritized and targeted using sensory grounding techniques to help the individual reliably regain contact with present external stimuli^97, 98^.

In addition to psychological interventions, some patients with highly distressing PTSD symptoms might respond to adjunctive pharmacological treatments such as Selective serotonin reuptake inhibitors (SSRIs) or other antidepressants blocking α^1^ adrenoceptors commonly used in patients with PTSD^99^. This integrated approach could be adapted depending on which mediators are present or predominant, as presented in **Figure 3**. For example, if there are no post-traumatic symptoms, the work on the schemas would be prioritised; similarly, the work on dissociative symptoms alone would be prioritised is these are predominant.

Thus, clinicians should carefully address subthreshold anxiety and depressive symptoms in traumatised individuals with psychosis. These could be, in isolation or in combination with other features (such as cognitive bias and posttraumatic symptoms) further targets for treatment (Target 3 **Figure 3**) that could indirectly ameliorate positive symptoms as suggested in previous reviews^12^. The use of antidepressants and CBT techniques such as behavioural activation and graded exposure^100^, as well as relaxation techniques and mindfulness, could then be useful, targeting low mood and anxiety respectively^100^.

Lastly, it is important to highlight the possible role of loneliness as another mediator that could be targeted for treatment. Promoting social inclusion and community membership through group interventions and vocational support could be beneficial, as well as improving existing interpersonal relationships using family therapy approaches. Also, as previously mentioned, loneliness might co-occur with other mediators.

In addition to pointing towards possible therapeutic interventions, our review indicates that routine assessment of trauma history should be necessary in clinical settings treating individuals with psychosis r Furthermore, given the possible importance of mediators, it is also important to carry out a careful assessment of the cognitive schemas, PTSD, dissociative symptoms, anxiety, mood and feelings of loneliness. These mediators can then be targeted through tailored psychological and/or pharmacological means, depending on which of them is predominant in the clinical picture.

## Conclusions

Our review suggests that the association between adversity and psychosis is mediating my various overlapping mechanisms. Cognitive schemas about the self and the world and post-traumatic symptoms (particularly dissociation) seem to play an important role in the association, while other factors such as mood and feeling of loneliness seem to contribute partially to this link and interact mutually in contributing to the effect. Our findings support the routine assessment of experiences of childhood adversity in clinical settings, alongside with all the potential mediators. An integrated approach targeting mediators such as mood or PTSD symptoms with psychological and/or medical means could potentially be a beneficial add-on to the traditional treatment of positive symptoms.

## Data Availability

This study is a systematic review and is based on data already published.

## References

1. Varese F, Smeets F, Drukker M, et al. Childhood adversities increase the risk of psychosis: a meta-analysis of patient-control, prospective-and cross-sectional cohort studies. Schizophrenia bulletin 2012;38(4):661–671.

2. Alameda L, Golay P, Baumann PS, Ferrari C, Do KQ, Conus P. Age at the time of exposure to trauma modulates the psychopathological profile in patients with early psychosis. J Clin Psychiatry May 2016;77(5):e612–618.

3. Ajnakina O, Trotta A, Oakley-Hannibal E, et al. Impact of childhood adversities on specific symptom dimensions in first-episode psychosis. Psychol Med Jan 2016;46(2):317–326.

4. Ruby E, Polito S, McMahon K, Gorovitz M, Corcoran C, Malaspina D. Pathways Associating Childhood Trauma to the Neurobiology of Schizophrenia. Frontiers in psychological and behavioral science Jan 1 2014;3(1):1–17.

5. Howes OD, McCutcheon R, Owen MJ, Murray RM. The Role of Genes, Stress, and Dopamine in the Development of Schizophrenia. Biological psychiatry Jan 1 2017;81(1):9–20.

6. Steullet P, Cabungcal JH, Monin A, Dwir D, O’Donnell P, Cuenod M, Do KQ. Redox dysregulation, neuroinflammation, and NMDA receptor hypofunction: A “central hub” in schizophrenia pathophysiology? Schizophr Res Sep 2016;176(1):41–51.

7. van Winkel R, van Nierop M, Myin-Germeys I, van Os J. Childhood trauma as a cause of psychosis: linking genes, psychology, and biology. The Canadian Journal of Psychiatry 2013;58(1):44–51.

8. Aas M, Haukvik UK, Djurovic S, et al. Interplay between childhood trauma and BDNF val66met variants on blood BDNF mRNA levels and on hippocampus subfields volumes in schizophrenia spectrum and bipolar disorders. Journal of psychiatric research 2014;59:14–21.

9. Teicher MH, Anderson CM, Polcari A. Childhood maltreatment is associated with reduced volume in the hippocampal subfields CA3, dentate gyrus, and subiculum. Proceedings of the National Academy of Sciences of the United States of America Feb 28 2012;109(9):E563–572.

10. Alameda L, Fournier M, Khadimallah I, et al. Redox dysregulation as a link between childhood trauma and psychopathological and neurocognitive profile in patients with early psychosis. Proceedings of the National Academy of Sciences 2018;115(49):12495–12500.

11. Myin-Germeys I, van Os J. Stress-reactivity in psychosis: evidence for an affective pathway to psychosis. Clinical psychology review 2007;27(4):409–424.

12. Bebbington P. Unravelling psychosis: psychosocial epidemiology, mechanism, and meaning. Shanghai Arch Psychiatry Apr 25 2015;27(2):70–81.

13. Morrison AP, Frame L, Larkin W. Relationships between trauma and psychosis: A review and integration. British Journal of Clinical Psychology 2003;42(4):331–353.

14. Garety PA, Bebbington P, Fowler D, Freeman D, Kuipers E. Implications for neurobiological research of cognitive models of psychosis: a theoretical paper. Psychological medicine 2007;37(10):1377–1391.

15. Gumley A, Taylor H, Schwannauer M, MacBeth A. A systematic review of attachment and psychosis: measurement, construct validity and outcomes. Acta Psychiatrica Scandinavica 2014;129(4):257–274.

16. Read J, Gumley A. Can attachment theory help explain the relationship between childhood adversity and psychosis? Attachment 2008;2(1):1–35.

17. Hardy A. Pathways from trauma to psychotic experiences: a theoretically informed model of posttraumatic stress in psychosis. Frontiers in psychology 2017;8:697.

18. Allen JG, Coyne L, Console DA. Dissociative detachment relates to psychotic symptoms and personality decompensation. Comprehensive psychiatry 1997;38(6):327–334.

19. Morgan C, Gayer□Anderson C. Childhood adversities and psychosis: evidence, challenges, implications. World Psychiatry 2016;15(2):93–102.

20. Read J, Fosse R, Moskowitz A, Perry B. The traumagenic neurodevelopmental model of psychosis revisited. Neuropsychiatry 2014;4(1):65–79.

21. Bentall RP, de Sousa P, Varese F, Wickham S, Sitko K, Haarmans M, Read J. From adversity to psychosis: pathways and mechanisms from specific adversities to specific symptoms. Social psychiatry and psychiatric epidemiology 2014;49(7):1011–1022.

22. Freeman D, Garety P. Advances in understanding and treating persecutory delusions: a review. Social psychiatry and psychiatric epidemiology 2014;49(8):1179–1189.

23. Misiak B, Krefft M, Bielawski T, Moustafa AA, Sąsiadek MM, Frydecka D. Toward a unified theory of childhood trauma and psychosis: a comprehensive review of epidemiological, clinical, neuropsychological and biological findings. Neuroscience & Biobehavioral Reviews 2017;75:393–406.

24. Williams J, Bucci S, Berry K, Varese F. Psychological mediators of the association between childhood adversities and psychosis: a systematic review. Clinical psychology review 2018;65:175–196.

25. Moher D, Liberati A, Tetzlaff J, Altman DG. Preferred reporting items for systematic reviews and meta-analyses: the PRISMA statement. Annals of internal medicine 2009;151(4):264–269.

26. Moher D, Booth A, Stewart L. How to reduce unnecessary duplication: use PROSPERO. BJOG: An International Journal of Obstetrics & Gynaecology 2014;121(7):784–786.

27. Hayes AF. Beyond Baron and Kenny: Statistical mediation analysis in the new millennium. Communication monographs 2009;76(4):408–420.

28. Diagnostic APA. statistical manual of mental disorders. American Psychiatric Association. Washington, DC 1994:886.

29. Organization WH. The ICD-10 classification of mental and behavioural disorders: clinical descriptions and diagnostic guidelines: Geneva: World Health Organization; 1992.

30. Van Os J, Linscott RJ, Myin-Germeys I, Delespaul P, Krabbendam L. A systematic review and meta-analysis of the psychosis continuum: evidence for a psychosis proneness–persistence–impairment model of psychotic disorder. Psychological medicine 2009;39(2):179–195.

31. Stang A. Critical evaluation of the Newcastle-Ottawa scale for the assessment of the quality of nonrandomized studies in meta-analyses. European journal of epidemiology 2010;25(9):603–605.

32. Alwin DF, Hauser RM. The decomposition of effects in path analysis. American sociological review 1975:37–47.

33. Evans GJ, Reid G, Preston P, Palmier-Claus J, Sellwood W. Trauma and psychosis: The mediating role of self-concept clarity and dissociation. Psychiatry Research 2015;228(3):626–632.

34. Morgan C, Reininghaus U, Fearon P, et al. Modelling the interplay between childhood and adult adversity in pathways to psychosis: initial evidence from the AESOP study. Psychological medicine 2014;44(2):407–419.

35. Peach N, Alvarez□Jimenez M, Cropper SJ, Sun P, Bendall S. Testing models of post□traumatic intrusions, trauma□related beliefs, hallucinations, and delusions in a first episode psychosis sample. British Journal of Clinical Psychology 2019;58(2):154–172.

36. Sun P, Alvarez-Jimenez M, Simpson K, Lawrence K, Peach N, Bendall S. Does dissociation mediate the relationship between childhood trauma and hallucinations, delusions in first episode psychosis? Comprehensive psychiatry 2018;84:68–74.

37. Appiah-Kusi E, Fisher H, Petros N, Wilson R, Mondelli V, Garety P, Mcguire P, Bhattacharyya S. Do cognitive schema mediate the association between childhood trauma and being at ultra-high risk for psychosis? Journal of psychiatric research 2017;88:89–96.

38. McDonnell J, Stahl D, Day F, McGuire P, Valmaggia L. Interpersonal sensitivity in those at clinical high risk for psychosis mediates the association between childhood bullying victimisation and paranoid ideation: a virtual reality study. Schizophrenia research 2018;192:89–95.

39. Thompson A, Marwaha S, Nelson B, Wood SJ, McGorry PD, Yung AR, Lin A. Do affective or dissociative symptoms mediate the association between childhood sexual trauma and transition to psychosis in an ultra-high risk cohort? Psychiatry research 2016;236:182–185.

40. Quidé Y, O’Reilly N, Watkeys O, Carr V, Green M. Effects of childhood trauma on left inferior frontal gyrus function during response inhibition across psychotic disorders. Psychological medicine 2018;48(9):1454–1463.

41. Cancel A, Comte M, Truillet R, et al. Childhood neglect predicts disorganization in schizophrenia through grey matter decrease in dorsolateral prefrontal cortex. Acta Psychiatrica Scandinavica 2015.

42. Chatziioannidis S, Andreou C, Agorastos A, Kaprinis S, Malliaris Y, Garyfallos G, Bozikas VP. The role of attachment anxiety in the relationship between childhood trauma and schizophrenia-spectrum psychosis. Psychiatry research 2019;276:223–231.

43. Choi JY, Choi YM, Kim B, Lee DW, Gim MS, Park SH. The effects of childhood abuse on self-reported psychotic symptoms in severe mental illness: Mediating effects of posttraumatic stress symptoms. 2015.

44. Hardy A, Emsley R, Freeman D, Bebbington P, Garety PA, Kuipers EE, Dunn G, Fowler D. Psychological Mechanisms Mediating Effects between Trauma and Psychotic Symptoms: The Role of Affect Regulation, Intrusive Trauma Memory, Beliefs, and Depression. 2016.

45. Isvoranu A-M, van Borkulo CD, Boyette L-L, Wigman JT, Vinkers CH, Borsboom D. A network approach to psychosis: Pathways between childhood trauma and psychotic symptoms. Schizophrenia Bulletin 2017.

46. Perona-Garcelan S, Carrascoso-Lopez F, Garcia-Montes JM, Ductor-Recuerda MJ, Jimenez AML, Vallina-Fernandez O, Perez-Alvarez M, Gomez-Gomez MT. Dissociative experiences as mediators between childhood trauma and auditory hallucinations. Journal of Traumatic Stress 2012.

47. Schalinski I, Breinlinger S, Hirt V, Teicher MH, Odenwald M, Rockstroh B. Environmental adversities and psychotic symptoms: The impact of timing of trauma, abuse, and neglect. 2019.

48. Steenkamp L, Weijers J, Gerrmann J, Eurelings-Bontekoe E, Selten J-P. The relationship between childhood abuse and severity of psychosis is mediated by loneliness: an experience sampling study. Schizophrenia research 2019.

49. Styła R, Stolarski M, Szymanowska A. Linking childhood adversities with schizophrenia: A mediating role of the balanced time perspective. Schizophrenia research 2019.

50. van Dam D, Korver-Nieberg N, Velthorst E, Meijer C, de Haan L. Childhood maltreatment, adult attachment and psychotic symptomatology: A study in patients, siblings and controls. Social Psychiatry and Psychiatric Epidemiology: The International Journal for Research in Social and Genetic Epidemiology and Mental Health Services 2014.

51. Varese F, Barkus E, Bentall R. Dissociation mediates the relationship between childhood trauma and hallucination-proneness. Psychological Medicine 2012.

52. Weijers J, Fonagy P, Eurelings-Bontekoe E, Termorshuizen F, Viechtbauer W, Selten J. Mentalizing impairment as a mediator between reported childhood abuse and outcome in nonaffective psychotic disorder. Psychiatry Research 2018.

53. Wickham S, Bentall R. Are specific early-life adversities associated with specific symptoms of psychosis?: A patient study considering just world beliefs as a mediator. Journal of Nervous and Mental Disease 2016.

54. Ashford CD, Ashcroft K, Maguire N. Emotions, traits and negative beliefs as possible mediators in the relationship between childhood experiences of being bullied and paranoid thinking in a non-clinical sample. Journal of Experimental Psychopathology 2012;3(4):624–638.

55. Bortolon C, Seillé J, Raffard S. Exploration of trauma, dissociation, maladaptive schemas and auditory hallucinations in a French sample. Cognitive neuropsychiatry 2017;22(6):468–485.

56. Boyda D, McFeeters D. Childhood maltreatment and social functioning in adults with sub-clinical psychosis. Psychiatry research 2015;226(1):376–382.

57. Boyda D, McFeeters D, Dhingra K, Rhoden L. Childhood maltreatment and psychotic experiences: Exploring the specificity of early maladaptive schemas. Journal of clinical psychology 2018;74(12):2287–2301.

58. Cole CL, Newman-Taylor K, Kennedy F. Dissociation mediates the relationship between childhood maltreatment and subclinical psychosis. Journal of Trauma & Dissociation 2016;17(5):577–592.

59. Fisher HL, Appiah-Kusi E, Grant C. Anxiety and negative self-schemas mediate the association between childhood maltreatment and paranoia. Psychiatry research 2012;196(2-3):323–324.

60. Fisher HL, Schreier A, Zammit S, Maughan B, Munafò MR, Lewis G, Wolke D. Pathways between childhood victimization and psychosis-like symptoms in the ALSPAC birth cohort. Schizophrenia bulletin 2012;39(5):1045–1055.

61. Gawęda Ł, Göritz AS, Moritz S. Mediating role of aberrant salience and self-disturbances for the relationship between childhood trauma and psychotic-like experiences in the general population. Schizophrenia research 2019;206:149–156.

62. Gibson LE, Reeves LE, Cooper S, Olino TM, Ellman LM. Traumatic life event exposure and psychotic-like experiences: A multiple mediation model of cognitive-based mechanisms. Schizophrenia research 2019;205:15–22.

63. Goodall K, Rush R, Grünwald L, Darling S, Tiliopoulos N. Attachment as a partial mediator of the relationship between emotional abuse and schizotypy. Psychiatry research 2015;230(2):531–536.

64. Jaya ES, Ascone L, Lincoln TM. Social adversity and psychosis: the mediating role of cognitive vulnerability. Schizophrenia bulletin 2016;43(3):557–565.

65. Lincoln TM, Marin N, Jaya ES. Childhood trauma and psychotic experiences in a general population sample: a prospective study on the mediating role of emotion regulation. European Psychiatry 2017;42:111–119.

66. Marwaha S, Bebbington P. Mood as a mediator of the link between child sexual abuse and psychosis. Social psychiatry and psychiatric epidemiology 2015;50(4):661–663.

67. Marwaha S, Broome MR, Bebbington PE, Kuipers E, Freeman D. Mood instability and psychosis: analyses of British national survey data. Schizophrenia bulletin 2013;40(2):269–277.

68. McCarthy-Jones S. Post-traumatic symptomatology and compulsions as potential mediators of the relation between child sexual abuse and auditory verbal hallucinations. Behavioural and cognitive psychotherapy 2018;46(3):318–331.

69. Murphy S, Murphy J, Shevlin M. Negative evaluations of self and others, and peer victimization as mediators of the relationship between childhood adversity and psychotic experiences in adolescence: the moderating role of loneliness. British Journal of Clinical Psychology 2015;54(3):326–344.

70. Perona-Garcelán S, García-Montes JM, Rodríguez-Testal JF, et al. Relationship between childhood trauma, mindfulness, and dissociation in subjects with and without hallucination proneness. Journal of Trauma & Dissociation 2014;15(1):35–51.

71. Pinto□Gouveia J, Matos M, Castilho P, Xavier A. Differences between depression and paranoia: the role of emotional memories, shame and subordination. Clinical psychology & psychotherapy 2014;21(1):49–61.

72. Rössler W, Ajdacic-Gross V, Rodgers S, Haker H, Müller M. Childhood trauma as a risk factor for the onset of subclinical psychotic experiences: Exploring the mediating effect of stress sensitivity in a cross-sectional epidemiological community study. Schizophrenia research 2016;172(1-3):46–53.

73. Sheinbaum T, Kwapil TR, Barrantes-Vidal N. Fearful attachment mediates the association of childhood trauma with schizotypy and psychotic-like experiences. Psychiatry research 2014;220(1-2):691–693.

74. Shevlin M, McElroy E, Murphy J. Loneliness mediates the relationship between childhood trauma and adult psychopathology: evidence from the adult psychiatric morbidity survey. Social psychiatry and psychiatric epidemiology 2015;50(4):591–601.

75. Sitko K, Bentall RP, Shevlin M, Sellwood W. Associations between specific psychotic symptoms and specific childhood adversities are mediated by attachment styles: an analysis of the National Comorbidity Survey. Psychiatry research 2014;217(3):202–209.

76. van Nierop M, Van Os J, Gunther N, et al. Does social defeat mediate the association between childhood trauma and psychosis? Evidence from the NEMESISL2 S tudy. Acta Psychiatrica Scandinavica 2014;129(6):467–476.

77. Wolke D, Lereya S, Fisher H, Lewis G, Zammit S. Bullying in elementary school and psychotic experiences at 18 years: a longitudinal, population-based cohort study. Psychological medicine 2014;44(10):2199–2211.

78. Yamasaki S, Ando S, Koike S, et al. Dissociation mediates the relationship between peer victimization and hallucinatory experiences among early adolescents. Schizophrenia Research: Cognition 2016;4:18–23.

79. Bortolon C, Raffard S. Dissociation Mediates the Relationship Between Childhood Trauma and Experiences of Seeing Visions in a French Sample. The Journal of nervous and mental disease 2018;206(11):850–858.

80. Isvoranu A-M, van Borkulo CD, Boyette L-L, Wigman JT, Vinkers CH, Borsboom D, Investigators G. A network approach to psychosis: pathways between childhood trauma and psychotic symptoms. Schizophrenia bulletin 2016;43(1):187–196.

81. Schalinski I, Breinlinger S, Hirt V, Teicher MH, Odenwald M, Rockstroh B. Environmental adversities and psychotic symptoms: The impact of timing of trauma, abuse, and neglect. Schizophrenia research 2017.

82. Perona□Garcelán S, Carrascoso□López F, García□Montes JM, Ductor□Recuerda MJ, López Jiménez AM, Vallina□Fernández O, Pérez□Álvarez M, Gómez□Gómez MT. Dissociative experiences as mediators between childhood trauma and auditory hallucinations. Journal of Traumatic Stress 2012;25(3):323–329.

83. Varese F, Barkus E, Bentall R. Dissociation mediates the relationship between childhood trauma and hallucination-proneness. Psychological medicine 2012;42(5):1025–1036.

84. Thompson A, Sullivan S, Lewis G, et al. Association between locus of control in childhood and psychotic symptoms in early adolescence: results from a large birth cohort. Cognitive neuropsychiatry 2011-Sep 2011;16(5):385–402.

85. Van Dam D, Korver-Nieberg N, Velthorst E, Meijer C, de Haan L. Childhood maltreatment, adult attachment and psychotic symptomatology: a study in patients, siblings and controls. Social psychiatry and psychiatric epidemiology 2014;49(11):1759–1767.

86. Wickham S, Bentall R. Are specific early-life adversities associated with specific symptoms of psychosis?: A patient study considering just world beliefs as a mediator. The Journal of nervous and mental disease 2016;204(8):606.

87. Hardy A, Emsley R, Freeman D, Bebbington P, Garety PA, Kuipers EE, Dunn G, Fowler D. Psychological mechanisms mediating effects between trauma and psychotic symptoms: the role of affect regulation, intrusive trauma memory, beliefs, and depression. Schizophrenia Bulletin 2016;42(suppl_1):S34–S43.

88. Weijers J, Fonagy P, Eurelings-Bontekoe E, Termorshuizen F, Viechtbauer W, Selten J. Mentalizing impairment as a mediator between reported childhood abuse and outcome in nonaffective psychotic disorder. Psychiatry research 2018;259:463–469.

89. Choi JY, Choi YM, Kim B, Lee DW, Gim MS, Park SH. The effects of childhood abuse on self-reported psychotic symptoms in severe mental illness: Mediating effects of posttraumatic stress symptoms. Psychiatry research 2015;229(1-2):389–393.

90. Moskowitz A, Schäfer I, Dorahy MJ. Psychosis, trauma and dissociation. Emerging Perspectives on Severe Psychopathology 2008.

91. Ross CA. Dissociation and psychosis: The need for integration of theory and practice. 2006.

92. Sin J, Spain D. Psychological interventions for trauma in individuals who have psychosis: a systematic review and meta-analysis. Psychosis 2017;9(1):67–81.

93. Swan S, Keen N, Reynolds N, Onwumere J. Psychological interventions for post-traumatic stress symptoms in psychosis: a systematic review of outcomes. Frontiers in psychology 2017;8:341.

94. Brand RM, McEnery C, Rossell S, Bendall S, Thomas N. Do trauma-focussed psychological interventions have an effect on psychotic symptoms? A systematic review and meta-analysis. Schizophrenia research 2018;195:13–22.

95. Garety PA, Kuipers E, Fowler D, Freeman D, Bebbington P. A cognitive model of the positive symptoms of psychosis. Psychological medicine 2001;31(2):189–195.

96. Howes OD, Murray RM. Schizophrenia: an integrated sociodevelopmental-cognitive model. The Lancet 2014;383(9929):1677–1687.

97. Keen N, Hunter E, Peters E. Integrated trauma-focused cognitive-behavioural therapy for post-traumatic stress and psychotic symptoms: a case-series study using imaginal reprocessing strategies. Frontiers in psychiatry 2017;8:92.

98. Steel C, Hardy A, Smith B, et al. Cognitive–behaviour therapy for post-traumatic stress in schizophrenia. A randomized controlled trial. Psychological Medicine 2017;47(1):43–51.

99. Steckler T, Risbrough V. Pharmacological treatment of PTSD–established and new approaches. Neuropharmacology 2012;62(2):617–627.

100. Waller H, Garety P, Jolley S, et al. Low intensity cognitive behavioural therapy for psychosis: a pilot study. Journal of behavior therapy and experimental psychiatry 2013;44(1):98–104.

